# General Practitioner perspectives and wellbeing during the COVID-19 Pandemic: a mixed method social media analysis

**DOI:** 10.1101/2021.10.19.21265194

**Authors:** Su Golder, Laura Jefferson, Elizabeth McHugh, Holly Essex, Claire Heathcote, Ana Castro Avila, Veronica Dale, Christina Van Der Feltz-Cornelis, Karen Bloor

## Abstract

**Background:** General practitioners (GPs) adapted their work practices rapidly in response to the COVID-19 pandemic. Limited research has explored their perspectives over this time, and factors that may affect their wellbeing.

**Method:** We conducted a social media analysis of NHS GPs practising in the UK during the COVID-19 pandemic to identify issues which may affect their wellbeing. To identify trends, we assessed 91,034 tweets from 185 GPs on Twitter who posted before and during the pandemic, (January 2019 to February 2021). To identify themes related to wellbeing, we analysed qualitatively 7145 tweets posted during the pandemic from 200 GPs.

**Results:** We identified inter-connecting themes that affect GP wellbeing, predominately around resources and support. Lack of personal protective equipment (PPE) and testing led to discussion of safety and risk, as well as increased workload resulting from staff isolating. Expressions of low morale and feeling undervalued were widespread, resulting from the perceived lack of support from the government, media and the general public at a time of staff shortages and high workload.

Trends in themes were apparent, with emphasis on PPE, testing and safety March to May 2020 and morale, abuse, ‘closed’ GP surgeries, testing, flu vaccines and overworked September to October 2020. From December 2020 the COVID-19 vaccine dominated posts.

**Conclusion:** GPs’ experiences and perceptions as reflected in their social media posts during the pandemic have changed over time; perceived lack of support and resources, and negative public perceptions have exacerbated their concerns about existing underlying pressures.

## Introduction

Before the COVID-19 pandemic, rising demands on UK NHS general practitioners (GPs) including increasing complexity and intensity of work and difficulties in recruitment and retention led to reports of a service in ‘crisis’ [1] and a ‘fundamental threat to the sustainability of primary care in England’ [2]. Forty per cent of NHS doctors reported psychological and emotional conditions before the pandemic [3], with GPs in particular experiencing high levels of burnout [4, 5].

COVID-19 brought about many adaptations to general practice, including a rapid transition to e-consultations, creation of ‘hot’ and ‘cold’ clinics (those examining patients experiencing COVID-19 symptoms and those addressing unrelated needs), managing the manifestations of the pandemic in their practice, managing risk, and responding to rapidly evolving guidelines. Indeed, high levels of stress and burnout amongst GPs during the COVID19 pandemic have been reported internationally, with an expanding literature on this topic (e.g. [6-8]. Two UK studies of GP pandemic experiences show greater work stress due to rapid change and uncertainty [9] and the importance of teams in creating a sense of solidarity [10]. These studies are relatively small - Trivedi [9] studied 111 GPs in Leicestershire and Wanat et al [10] is a cross-European qualitative interview study of primary care staff, but includes only seven GPs in England. Research studying the pandemic experiences of a national sample of UK GPs is needed.

Social media use by health professionals has become increasingly widespread [11, 12] and this has expanded during the pandemic; Twitter is one of the most common types of social media platform [13-15]. Increasing numbers of GPs are using social media, and increasing frequency of posts provides a useful tool for ascertaining their personal views and experiences. Doctors use Twitter to discuss, rapidly and informally, current issues pertinent to their work and to communicate with colleagues [13]. Unsurprisingly, this has focused recently on issues related to COVID-19 and work practices [16]. Analysis of Twitter posts is used commonly to identify public experiences and opinions, including recently on COVID-19 and its impact [17-31].Medical professionals’ opinions about COVID-19 have been studied using social media [32], but no studies to date have examined GP wellbeing using social media monitoring, which is the aim of this study.

## Methods

Firstly, we sought to explore trends pre- and post-COVID using a longitudinal analysis of GPs’ tweet content (January 2019 to February 2021). Secondly, a more qualitative analysis explored themes emerging from GP tweets during the pandemic (February 2020 to February 2021) with a particular focus on GP wellbeing. This analysis was inductive in nature, and as such we did not seek to confirm or refute an existing hypothesis, but rather to explore emerging themes. This paper conforms to the Standards for Reporting Qualitative Research (SRQR).

### Sampling and data

Twitter profiles of users who tweeted on COVID-19 from 10 March 2020 to October 2020 were shared by Professor Mike Thelwall (see acknowledgments). We limited profiles to those with a self-reported UK location and ‘Dr’ or ‘Doctor’ in their username (5,512 users and 223 users, respectively) or biographical description (850 users and 3,885 users). We then selected GPs manually, excluding non-NHS GPs, retired GPs and practice or organisation accounts. This identified 293 practising UK NHS GPs. We supplemented these by searching for ‘NHS GP’ as a phrase in ‘people’ in the Twitter Advanced Search facility, identifying a further 88 Twitter users after removing duplicates. The resulting sample included 381 UK NHS GPs.

To explore the representativeness of our Twitter sample, we collated available demographic data, such as gender and race (categorised as black, white or Asian), geographical location, and type of GP (such as GP partner or GP trainee).

The longitudinal analysis included 185 GPs from the total 381 sample, those for whom we could obtain continual tweets from 1^st^ January 2019 through to February 2021. This number was a subset due to a number of factors. A Twitter download limit of 3200 tweets per user prevented some access to tweets from January 2019 for very high users. In addition, some GPs joined after January 2019, had an extended break from Twitter, or changed their account to private.

For the qualitative analysis one author (SG) randomly selected 200 GPs from the total 381 who posted tweets from February 2020. This generated variation in demographics and posts, whilst enabling data saturation with no new themes emerging. Of these 200 GPs, 196 had timelines containing data relevant to GP wellbeing and were included in our analysis.

### Analysis

For both analyses we collected tweets from the GPs sampled using Mozdeh software (http://mozdeh.wlv.ac.uk/) excluding non-English tweets, retweets, and duplicate tweets and imported them into an Excel Spreadsheet. The longitudinal analysis analysed trends over time in #hashtags, @handles/usernames, words used and key themes. The number of occurrences may disproportionately reflect use by a few prolific GPs, so we also recorded mentions by overall number of GPs.

In the qualitative analysis SG, LJ, EM, HE, CH, AC (university health researchers with experience in qualitative research) pursued a more in-depth manual content analysis exploring themes emerging during the pandemic. Content analysis involves coding sets of texts (tweets) into multiple relevant categories [31]. It is one of the most common methods for studying information obtained from social media [33] and is appropriate for identifying prevalence [31]. We used an inductive approach as we were not testing an existing theory and had no prior framework.

To answer our research questions fully, data immersion was essential. Following data familiarisation and immersion, a coding framework and annotation guide was developed by SG, then discussed and refined through multiple iterations with the study team. Tweets were hand-coded as this is still the gold standard form of analysis [34]. To avoid over-interpretation of these short tweets, we coded only what was explicitly stated. We tested for consistency in coding to increase the dependability of the findings by independently double coding 10% of tweets. Level of agreement was high, with 1.2% (11/915) codes changed and three additional codes added to existing codes. Remaining codes were checked by the second reviewer during the coding categorisation process, rather than independently assigned.

## Results

### Sample demographics

Our sample reflects the GP population in terms of broad ethnic group, but over-represents men (Table 1). GPs were located throughout the UK, with a slight over-representation from London. The majority (81%) did not indicate what GP role they held. Of those that did, most were GP Partners or GP Trainees/Registrars. Age was reported by less than 5% of GPs.

**Table 1:** GP Demographics (Gender, Race and Country)

|  | Male | Female | Unknown | White | Asian | Black | Unknown | England | Scotland | Wales | NI | Unknown UK |
| --- | --- | --- | --- | --- | --- | --- | --- | --- | --- | --- | --- | --- |
| Longitudinal Study (n=185) | 58% (107/185) | 42% (77/185) | 0.5% (1/185) | 64% (118/185) | 30% (55/185) | 4% (7/185) | 3% (5/185) | 75% (138/185) (35 from London) | 6% (11/185) | 4% (8/185) | 2% (3/185) | 14% (25/185) |
| Qualitative Study (n=196) | 56% (109/196) | 43% (85/196) | 1% (2/196) | 61% (119/196) | 32% (63/196) | 6% (11/196) | 2% (3/185) | 77% (150/196) (36 from London) | 7% (13/196) | 5% (9/196) | 1% (2/196) | 11% (22/196) |
| GP population (Headcount)* | 43% | 56% | 1% | 51% | 23% | 4% | 19% | 82% | 9% | 6% | 3% | - |

\*Gender and ethnicity is for England
NB: Data is from - England https://digital.nhs.uk/data-and-information/publications/statistical/general-practice-workforce-archive/31-march-2021, Wales https://gov.wales/general-practice-workforce-30-september-2020, NI http://www.hscboard.hscni.net/our-work/integrated-care/gps/gps-current-facts-and-figures/, and Scotland https://publichealthscotland.scot/publications/general-practice-gp-workforce-and-practice-list-sizes/general-practice-gp-workforce-and-practice-list-sizes-2010-2020/.

### Analysis 1: Longitudinal Trends January 2019 to February 2021

#### Volume of Tweets (91,034 tweets from 185 GPs)

The number of tweets increased dramatically a few days before the first UK lockdown (20^th^ March 2020) and stayed high until the end of April 2020 (Figure 1). The next largest peaks were around the time of the Royal College of General Practitioners (RCGP) Annual Conference (24^th^-25^th^ October 2019) and the US presidential election results (November 2020). Other smaller peaks reflected polling day (12^th^ December 2019), a GP conference “DGPLondon20” (29^th^ February 2020), announcement of the second lockdown (31^st^ October 2020), the first vaccine efficacy results (12^th^ November 2020), and the COVID-19 vaccine roll-out (January 2021).

**Figure 1:**
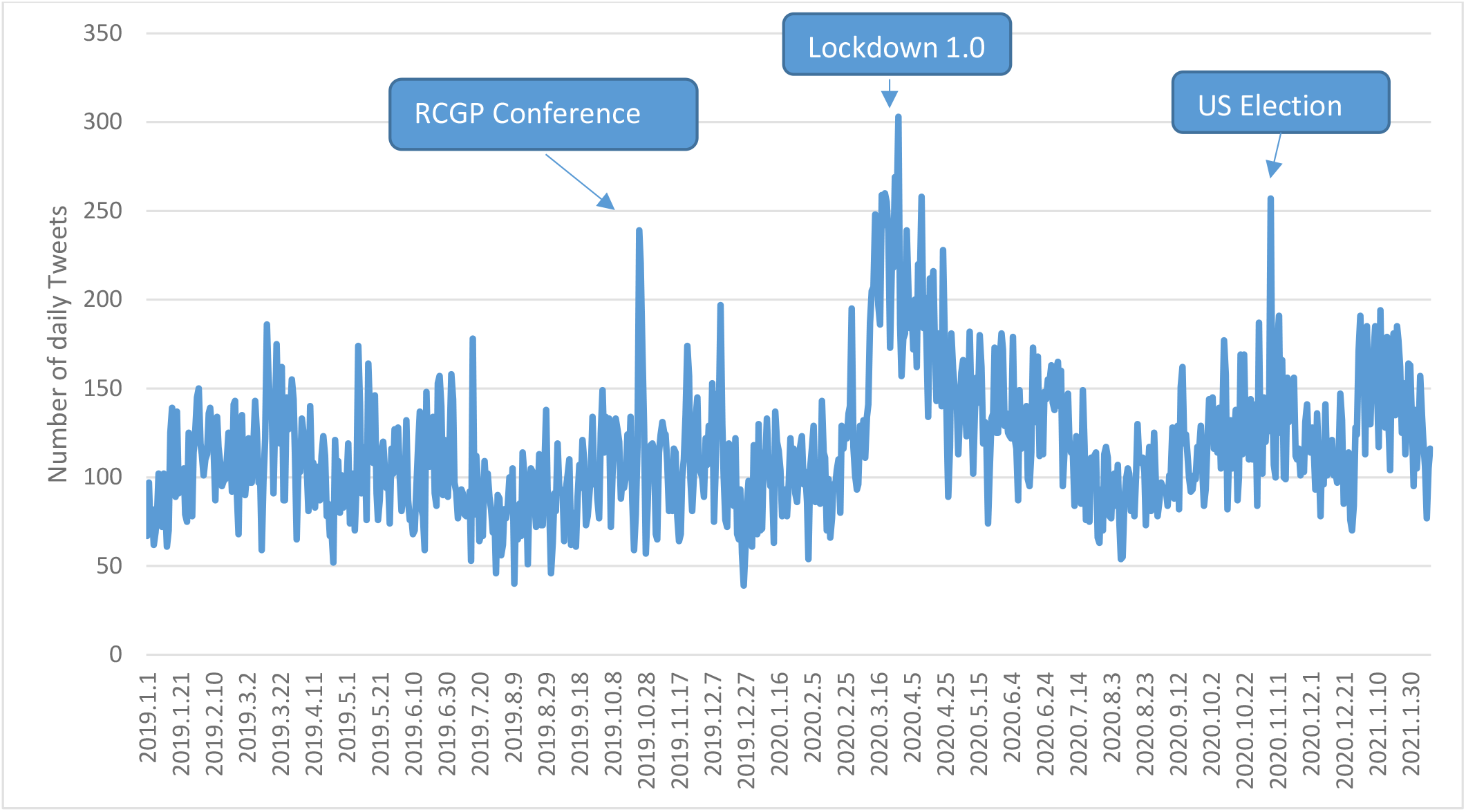
Total number of daily Tweets from 185 GPs.

#### Hashtags (11,950 unique hashtags mentioned 34,372 times)

In addition to hashtags related to the NHS and primary care, which dominate GPs’ tweets throughout the time period, those related to Brexit dominated in 2019, COVID-19 dominated in 2020 and COVID-19 vaccines in 2021 (Supplementary Table 1). Many new COVID-19 hashtags emerged in 2020, such as, #covid19, #coronavirus, #stayhomesavelives, #lockdown, #socialdistancing, #covidvaccine, #ppe, and #nhsheroes.

#### Handles (34,931 unique handles used 177,766 times)

The most common Twitter handles cited by GPs were organisations (such as @rcgp, @nhsengland, @thebma), politicians (such as @matthancock, @borisjohnston) and fellow GPs. Handles in 2020/2021 were similar to 2019 with an increase in mentions of GPs labelled as ‘renowned COVID-19 health experts’ by Twitter (Supplementary Table 2).

#### Words (86,671 different words used 1,731,115 times)

Similar language was used in each year (Supplementary Table 3). Many of the words used were to thank the hard work of colleagues (thank, time, great, work, staff, team, practice). Terms in 2021 reflected the COVID-19 vaccine rollout.

#### Specified Themes (using words and hashtags)

During the first wave, tweets related to COVID-19 and interventions to reduce transmission (such as lockdown, social distancing and PPE) were common, as were issues around safety, frontline staff and mortality (Supplementary Table 4 and Figure 2). Lack of testing tweets peaked in both the first and second wave. Commentary around remote working increased during the pandemic. References to workload, being closed, flu, GP ‘bashing’ and low morale peaked around September to October 2020. Issues related to coping, wellbeing, appreciation, sadness and enjoyment appeared throughout the time period. January 2021 saw an increase in vaccine-related tweets.

**Figure 2:**
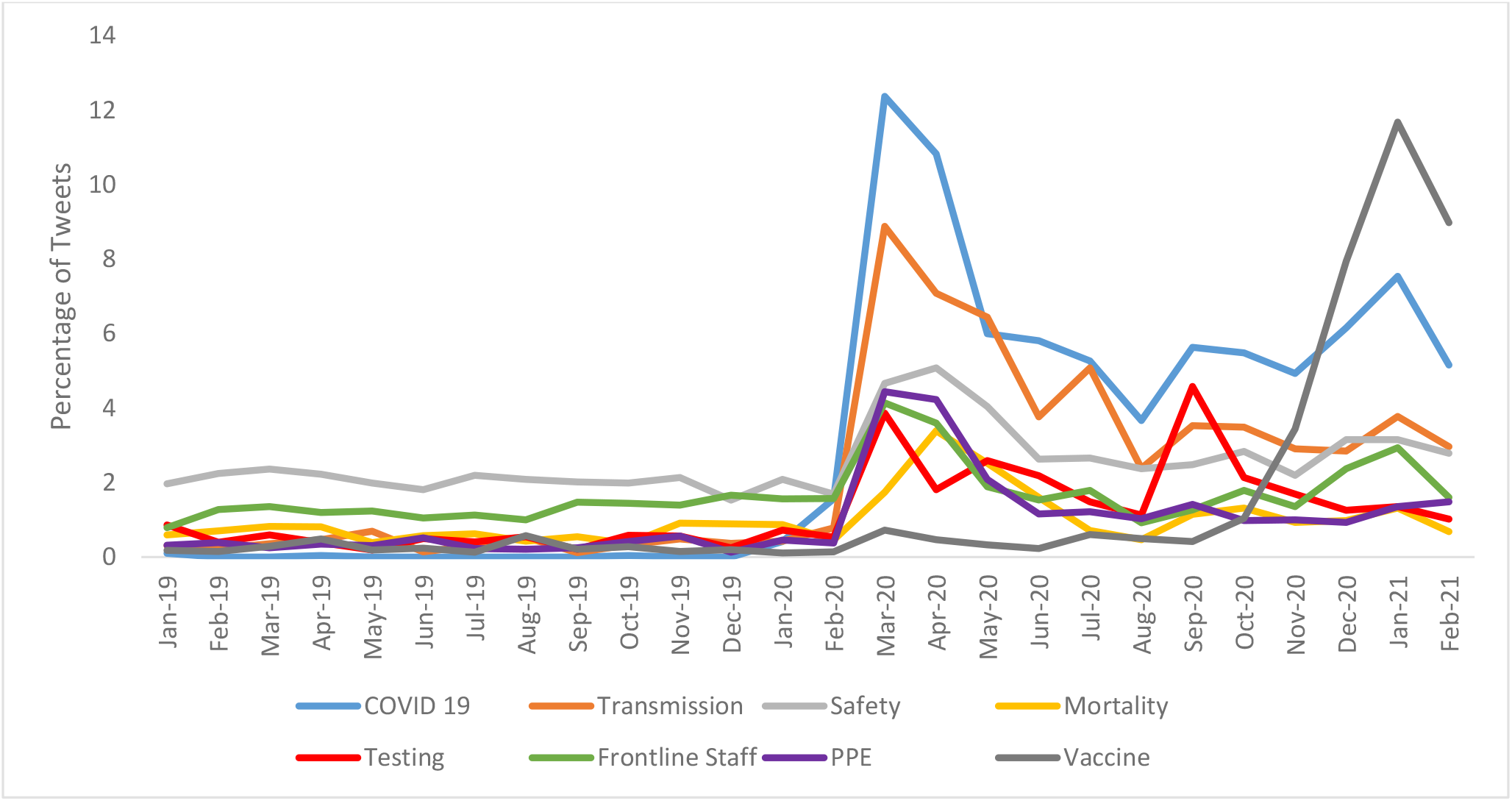
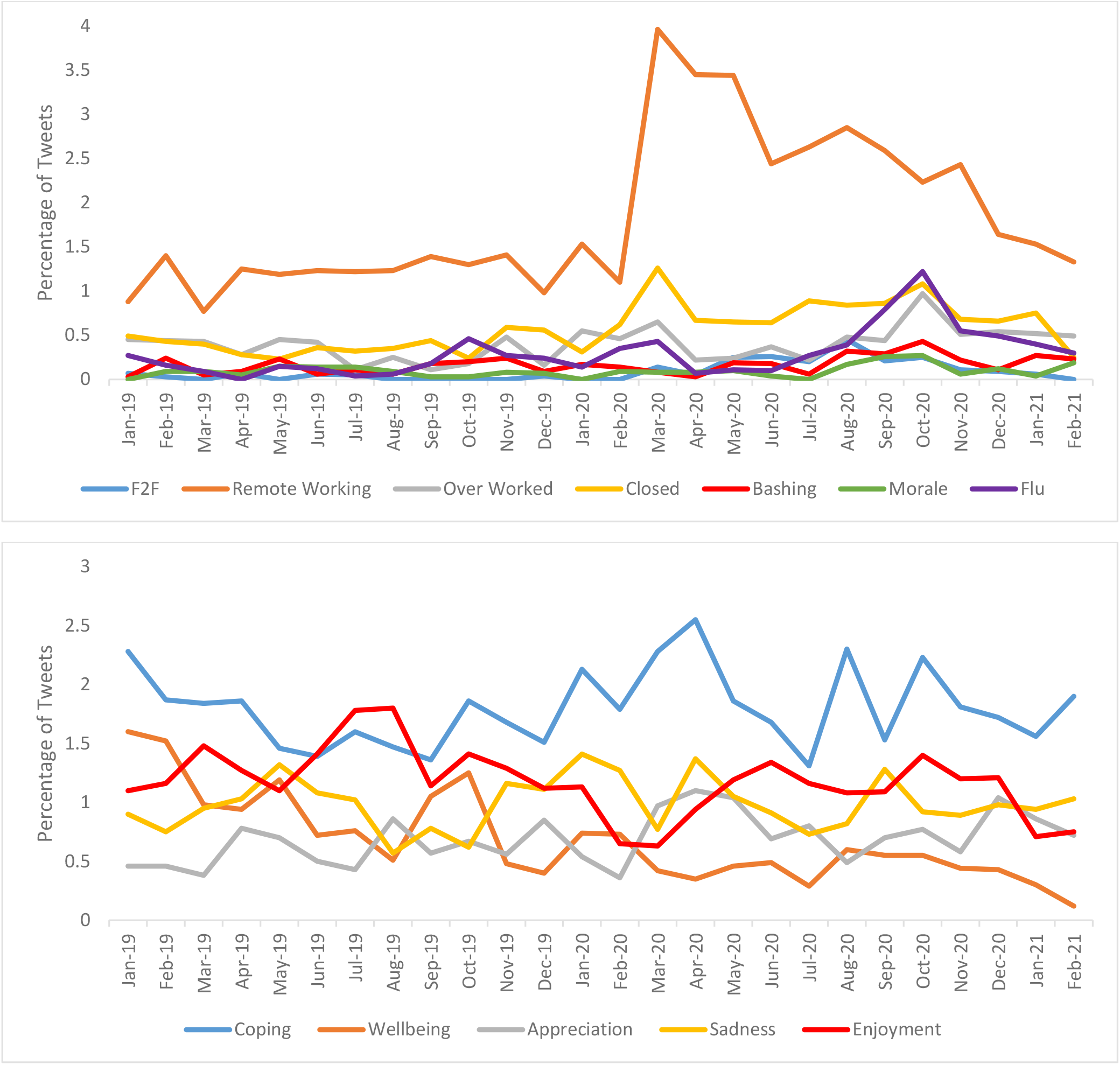
Daily percentages of GP tweets mentioning specific related-terms.

### Analysis 2: Qualitative exploration

Table 2 summarises the themes which emerged during the pandemic. The 12 most identified themes in descending order of frequency are described. Some of the themes are interconnected.

**Table 2:** Main themes and sub-themes of qualitative content analysis.

| Themes | Sub-themes | No of tweets (n=7145*) | No of GPs (n=196) | % of tweets made by female, male GPs (n=86, 110)** | % of tweets posted by White, Asian, Black GPs (n=119, 63, 11)*** |
| --- | --- | --- | --- | --- | --- |
| Changes to GP practice (n=1746, 24%) | Changes to practice (negative) | 946 (13%) | 142 (72%) | 64%, 78% | 71%, 75%, 64% |
|  | Changes to practice (positive or neutral) | 800 (11%) | 132 (67%) | 64%, 78% | 67%, 68%, 55% |
| NHS resources (n=1277, 18%) | Resources, lack of (general) | 118 (2%) | 42 (21%) | 12%, 28% | 18%, 22%, 27% |
|  | Resources, lack of (PPE) | 521 (7%) | 107 (55%) | 43%, 61% | 52%, 64%, 54% |
|  | Resources, lack of (testing) | 289 (4%) | 70 (36%) | 34%, 35% | 28%, 46%, 36% |
|  | Resources, lack of (staff) | 169 (2%) | 64 (33%) | 23%, 39% | 29%, 40%, 18% |
|  | Resources, lack of (funding, pay) | 141 (2%) | 47 (24%) | 20%, 26% | 20%, 30%, 27% |
|  | Resources, adequate | 39 (1%) | 26 (13%) | 6%, 19% | 9%, 17%, 27% |
| Direction/Management/Leadership from UK government or leading organizations such as RCGPs and BMA (n=1161, 16%) | Direction, management (positive) | 115 (2%) | 53 (27%) | 19%, 34% | 61%, 62%, 45% |
|  | Direction, management (negative) | 1046 (15%) | 119 (61%) | 56%, 63% | 26%, 30%, 18% |
| Information (n=1037, 15%) | Misinformation (about or received by GPs) | 564 (8%) | 109 (56%) | 57%, 54% | 54%, 57%, 64% |
|  | Information use and sharing (among GPs) | 343 (5%) | 89 (45%) | 44%, 45% | 39%, 52%, 64% |
|  | Information to support GP wellbeing | 130 (2%) | 53 (27%) | 30%, 24% | 23%, 33%, 27% |
| Appreciation of or by GPs (n=1015, 14%) | Appreciation of GPs (negative) | 277 (4%) | 84 (43%) | 41%, 44% | 39%, 52%, 27% |
|  | Appreciation of GPs (positive) | 188 (3%) | 84 (43%) | 43%, 43% | 42%, 44%, 45% |
|  | Appreciation of others | 550 (8%) | 111 (57%) | 63%, 52% | 55%, 59%, 73% |
| Part of NHS (n=686, 10%) | NHS work, colleagues (positive) | 620 (9%) | 119 (61%) | 55%, 54% | 56%, 68%, 64% |
|  | NHS work, colleagues (negative) | 66 (1%) | 30 (15%) | 10%, 18% | 12%, 17%, 36% |
| Personal GP experiences or emotions (n=613, 9%) | Experience C19 positive test/self-isolation | 151 (2%) | 58 (30%) | 29%, 30% | 22%, 46%, 27% |
|  | Experience stress/burnout | 98 (1%) | 43 (22%) | 22%, 22% | 21%, 24%, 18% |
|  | Emotions | 364 (5%) | 103 (53%) | 64%, 43% | 53%, 49%, 55% |
| GP workload (n=552, 8%) | Workload increase | 537 (8%) | 105 (54%) | 45%, 59% | 52%, 57%, 45% |
|  | Workload decrease | 15 (0.2%) | 13 (7%) | 8%, 5% | 7%, 8%, 0% |
| Colleagues health or wellbeing (n=533, 7%) | Concern about colleague health, wellbeing (depression, burnout, etc.) | 533 (7%) | 120 (61%) | 59%, 62% | 51%, 75%, 91% |
| Risks to GPs (n=481, 7%) | Risks to GPs themselves | 367 (5%) | 89 (45%) | 43%, 46% | 39%, 56%, 37% |
|  | Risks to GPs families | 45 (1%) | 24 (12%) | 13%, 12% | 9%, 17%, 9% |
|  | Risks to Black, Asian and minority ethnic (BAME) GPs | 69 (1%) | 25 (13%) | 12%, 13% | 5%, 36%, 22% |
| Communication/integration/collaboration (n=294, 4%) | Communication (positive) | 208 (3%) | 69 (35%) | 29%, 40% | 34%, 37%, 27% |
|  | Communication (negative) | 86 (1%) | 44 (22%) | 16%, 27% | 21%, 25%, 18% |
| Self-care of GPs in reference to their wellbeing (n=201, 3%) | Self-care (positive) | 189 (3%) | 65 (33%) | 37%, 30% | 30%, 37%, 36% |
|  | Self-care (negative) | 12 (0.2%) | 9 (5%) | 3%, 5% | 5%, 3%, 9% |

\*Some tweets discussed more than one topic and were coded in more than one category.
\*\*Using test of proportions, comparing the % of male tweeting the topic and the % of female -topics of 'negative changes to practice' (p=0.028), 'lack of resources' (p=0.005), 'lack of staff' (p=0.019), 'PPE' (p=0.013), 'adequate resources' (p<0.001) and 'positive direction/management' (p=0.019) were posted more heavily by males than females, whereas experience of 'emotions' (p=0.003) were more by females than males.
\*\*\*Using Fisher's exact test significant differences were found in four categories about 'Risks to BAME GPs' (p<0.001), 'Resources lack of (testing)' (p=0.048), 'Experience C19' (p=0.003) and 'Colleague Health/Wellbeing' (p=0.001) with a higher proportion of Black and Asian GPs tweeting.

#### Changes to practice

Most posts regarding changes to practice were neutral or negative. Neutral posts simply stated changes being made, most commonly remote working (IT systems and software, working from home and triaging patients in their own homes) with some referring to safe practices for face-to-face consultations (cleaning between patients, PPE, scrubs, and ‘hot hubs’). Most of the negative comments about changing work practices related to remote working. GPs expressed concern around missed diagnoses, widening health inequalities and increased time and fatigue associated with remote consultations. GPs felt job satisfaction, personal care, emotional support and patient satisfaction were all challenged by remote working. Problems contacting patients were common because of patient availability, phone networks, internet providers, or IT systems. Working from home brought about additional challenges, especially for those with children.

GPs emphasized the need to remain accessible to patients through face-to-face consultations, but safety measures (e.g. PPE, cleaning) increased time pressures. Safely visiting care homes was a major challenge, with concerns around transmission to vulnerable patients. Challenges around patient non-attendance for potential cancer, stroke and heart attacks also caused anxiety for GPs, and they described an increasing number of patients with mental health problems.

There was concern about GPs delivering the COVID vaccination programme; some expressed the view that this could not be done without deprioritising other services, that reimbursement was low and that other health care professionals may be more appropriate as GP staffing levels were already critical.

There were a small number of positive posts praising colleagues or software or stating the benefits of remote consultations in terms of efficiency and patient care, alongside some speculation on whether some changes could be permanent by taking the ‘best of the changes into the future’. A few mentioned how retired GPs could be deployed by remote working.

Those with a preference for working from home remarked on greater work/life balance and extra time for housework or cooking. GPs appeared buoyed by the vaccine rollout, expressing positivity around their role in helping to ‘protect the UK population’ and a morale boost from patients’ feedback when receiving vaccinations.

#### Lack of Resources

##### COVID-19 Testing

GPs expressed anger about the lack of testing in the first phase of the pandemic, resulting in risk to patients and unnecessary self-isolation creating added staffing pressures. There was frustration that GPs were perceived to be lower priority than high-profile public figures, while ‘we risk our lives’. Furthermore, when comparing themselves to hospital staff, GPs were confused and angry as to why they were not a priority group, given they had more contact with patients.

GPs reported that the testing system again caused problems in September 2020 with long waiting times for results, or long distances to testing centres. GPs also complained about ‘rationing’ patient tests. Problems with the supply and accuracy of lateral flow tests was emphasised in January 2021.

##### Staff

Reported concerns related to a perceived shortage of doctors and nurses, with many referencing a decline in GP numbers over recent years, expressing the view that primary care staffing levels were critical before the pandemic and COVID-19 self-isolation exacerbated these problems. GPs reported attrition throughout the pandemic, due to factors such as workload, underfunding, low morale and lack of appreciation.

##### Personal Protective Equipment (PPE)

Lack of PPE appeared to be a common problem leading some GPs to report purchasing their own, reusing equipment, improvising, accepting donations or posting pleas on social media for supplies. GPs felt they were lower priority than secondary care staff and ‘even supermarket employees’, and initial public panic buying meant low supplies of hand sanitiser and cleaning products.

The quality of PPE was described as ‘substandard’ or even ‘hopeless’ with flimsy paper masks, thin plastic aprons and masks four years out of date. Pleas were made for World Health Organization guidance on PPE to be followed including FFP3 masks. Questions on the effectiveness of surgical masks increased as more emphasis was placed on aerosol transmission.

There were concerns expressed about being silenced, with social media appeals taken down, Clinical Commissioning Groups asking GPs not to speak out and media reports of ‘whistleblowers threatened with job loss for speaking out on PPE’. GPs expressed anger at the UK Health Secretary’s comment that PPE should be treated as a ‘precious resource’ as this appeared to blame staff for not using equipment carefully. GPs questioned how many lives were lost because of inadequate PPE supplies.

##### Funding and Pay

Many GPs posted about funding cuts over the past 10 years. The frustration surrounding perceived underfunding of primary care grew over time as increasing amounts were spent on privately provided interventions including ‘NHS Test and Trace’, which many GPs felt would have been better managed by the NHS.

There was reported concern over what was viewed as a real terms pay cut, and frustration when comparisons were made to MPs’ salaries or management consultants funded by government.

#### Adequate Resources

These posts were far fewer in number (see Table 2) and tended to refer to having PPE or testing available. Some of these stated that ‘at last we have plenty of PPE’, thus referring to a time when supplies were inadequate.

#### Direction and management

Approximately a tenth of posts relating to direction and management were positive, and these were mostly directed at organisations such as the RCGP, King’s Fund, or Public Health England with a few directed at government actions such as the NHS workers’ visa extension and abolishing NHS fees for overseas staff.

The vast majority of posts, though, were negative. A few GPs were negative about organisations such as the RCGP, NHS England and the BMA, expressing a need for more support and action. Most criticism was focused at the government, particularly in England. Issues were related to other themes in our analysis such as underfunding, declining numbers of GPs, lack of PPE and testing, inconsistent or poor guidance for GPs and GPs being used as a scapegoat.

Shielding lists were seen by some as poorly managed centrally, with GPs rectifying errors. The paperwork required for returning GPs was criticised, and some thought the scheme was putting older people at risk. Poor management of care homes during the pandemic and risks to staff and patients were also discussed; there were calls for the government to be accountable for the deaths of NHS staff.

GPs felt overlooked by government with regard to testing and mandatory public face coverings. There was a general sentiment that the focus of government and media was on hospital patients and staff. Anger was expressed about the behaviour of political figures who ‘break the rules’.

Some GPs did not feel properly supported to carry out the COVID-19 vaccination programme and held concerns around supply chain issues and fielding patient enquiries.

#### Misinformation about or received by GPs

Some GPs complained of confusing information and guidance received. For example, in March 2020, concerns were raised about the mask policy in primary care, when GPs should isolate, suspension of routine work, and whether to move to remote consultations. As the pandemic progressed other issues were raised such as the ‘shambolic’ correcting of shielding, advice from NHS 111 and government for the public to contact their GP inappropriately, and misinformation on dexamethasone use.

Misinformation about GP surgeries being ‘closed’ persisted through the pandemic, with peak frustration expressed by our sample of GPs in April and September 2020. The government was seen as perpetuating this misinformation. Many GPs were particularly affronted by a letter from NHS England in September 2020 to remind GP practices of their duty to provide face-to-face appointments, warning that failure to do so could constitute a breach of contract. GPs declared that they were not ‘tucked away safe’ or ‘twiddling our thumbs’ but ‘working harder than ever’. Pleas to the public and reassurances that practices were open continued into 2021.

#### Information use and sharing among GPs

Some GPs shared advice on working practices, whilst others asked questions of their colleagues. Sharing petitions on issues such as testing or PPE and work surveys was commonplace, as was sharing factual information on issues such as GP deaths, risks for BAME GPs, and doctors with long COVID.

#### Information to support GP wellbeing

These posts often consisted of links to webinars, events or resources to support GP wellbeing. Other posts simply suggested ways to help such as ‘being kind to yourself’, ‘taking breaks’, ‘taking down time’ and ‘keeping active’.

#### (Lack of) appreciation of GPs

Many GPs expressed feelings of being unappreciated by media, government and the public. In the first lockdown there were a few reports of stealing from GP practices such as toilet rolls and hand sanitizer as well as vandalism, graffiti and abuse from patients. Public perceptions were viewed as being exacerbated by GPs being accused of being ‘closed’, needing to ‘reopen’ and the insinuation that GPs are ‘sitting around doing nothing’, ‘lazy’, ‘selfish’ and not doing the job they ‘signed up to’. GPs expressed frustration about this and felt they experienced a lack of respect and appreciation. There were references to ‘GP bashing’ and GPs being blamed for the ‘failures of government’. GPs complained of feeling like ‘public enemy number one’, particularly from September 2020.

The ‘clap for carers’ was met with a positive emotional response by some GPs. Some said that ‘it bought a lump to my throat’ or ‘a tear to my eye’. As time went on, however, expressed views of this became less positive and by January 2021, discussion around a return of ‘clap for carers’ was met with calls for the public instead to observe the rules to protect the NHS.

There were fewer posts relating to positive appreciation of GPs and these tended to reference appreciation shown in the form of gifts from businesses or the local community, allocated shopping times, donations of PPE, and clapping in the first lockdown. GPs recognised patient gratitude, particularly during the vaccination programme.

#### Appreciation of others

GPs expressed gratitude to those supporting them in the pandemic, including organisations, volunteers, the public, neighbours, and local businesses. There was also admiration for other professions such as scientists and teachers.

#### NHS work colleagues

There were expressions of gratitude to NHS staff, particularly primary care colleagues, secondary and community care staff (particularly care homes), domestic staff and administrative teams in hospitals. There were comments about altruism and dedication with staff ‘going above and beyond the call of duty’, ‘showing courage’ and being ‘amazing’, ‘world class’ and ‘heroes’. There were also views expressed about how ‘fantastic’ the NHS itself is by providing free care, rapid adaptations to change and an incredible response to the crisis.

There was, however, some criticism of other parts of the NHS treating GPs ‘like commodities instead of human beings’, ‘bullying’, ‘too much bureaucracy’, and a management team ‘devoid of reality’.

#### Colleague health and wellbeing

Issues were raised regarding burnout, stress, anxiety and even suicide resulting from the ‘extreme pressure’ and ‘overwhelming workload’. There were concerns about GPs leaving the profession at ‘an alarming rate’, and calls for support for GP wellbeing. There was resistance to ‘resilience training’, seen by some as ‘blaming colleagues’ and thus ‘insulting’; others welcomed ‘support groups’, ‘retreats’ and being kind to colleagues.

GPs expressed worries for their colleagues’ safety, particularly given lack of PPE and testing, likening GPs to ‘soldiers fighting without armour’. GPs felt their colleagues were ‘putting their lives on the line’, particularly returning retired GPs and BAME GPs. In response to the risks, GPs reported that colleagues were ‘preparing their wills’, looking into death in service benefits or seeking guardianship of their children in preparation for the worst. Other posts announced colleagues hospitalised with or dying from COVID-19, and numbers of GPs dying.

#### Workload

Throughout the time-period under consideration, GPs expressed anxiety over their workload. This resulted in GPs reporting working long shifts, frequently working over 50 hour weeks, working on days off, not taking annual leave or bank holidays. Before COVID-19, primary care was described as at ‘breaking point’. During the pandemic, workload was described as having ‘gone through the roof’ creating ‘immense pressure’ with GPs ‘pushed to the limit’. The situation was described as unsustainable.

Additional pressures reported included the increase in remote consultations (described as taking longer), NHS 111 referrals, hospitals reducing non-COVID-19 services, keeping up to date with COVID-19 evidence and safe working practice guidelines, dealing with patient shielding lists, donning and doffing PPE, sanitising between patients, requests for mask exemption letters, shielding notes, isolation notes, sick notes, and early ordering of prescriptions. In addition, patient demand was perceived to have increased due to a rise in mental health issues, an expanded flu vaccination programme and the COVID-19 vaccination rollout.

A very small minority of posts related to a reduction in workload, mostly during April 2020 (see Table 2).

Discussion regarding ways in which workload could be managed centred around improving mental health services, community volunteers, self-referral services, increased capacity, and managing patient contact with practices for unnecessary reasons (e.g. vaccination dates). Some GPs reported working part-time as a means of coping with the workload.

#### Emotions and stress

GPs reported anxiety about their own safety, the safety of their families, the ‘tsunami’ in workload, and lack of resources. Many stated that they were ‘fatigued’ or ‘exhausted’, ‘fearful’ about the level of care for patients, and ‘heartbroken’ by patients suffering or dying alone. GPs talked about being ‘fearful about the uncertainties’, and ‘dreading the future’.

GPs also referred to the pressures of providing care in a pandemic, using phrases such as ‘unbearable pressure’, ‘completely overwhelmed’, and ‘never felt so exhausted’. Some perceived an impact on their mental health, with comments that they felt ‘mentally drained’, ‘broken’, ‘wiped out’, ‘worn down’, ‘teary’ and ‘burnt out’.

GPs reported feeling like ‘nobodies’ or ‘expendable’ and viewed this as resulting from government and media actions. Low morale was exacerbated by ‘false rumours’ and a ‘constant attack on GPs’ by media, government and the public. Many GPs commented on how they were feeling frustrated or ‘insulted’ by public behaviour such as noncompliance with lockdowns or isolation, not wearing masks, and vaccine uptake. Others mentioned feeling guilty because they were shielding or taking annual leave, or even feeling guilty for catching COVID-19 as if it was a ‘lifestyle choice’. Some felt ‘not listened to’ and powerless in the face of adversity.

From late November there were more positive posts with GPs reporting that they were at last ‘feeling hopeful’. Reports of getting the COVID-19 vaccine were met with comments like ‘Fantastic Day!’, ‘feeling privileged’ and ‘so delighted’. Those reporting on their involvement with the vaccine roll-out described feeling ‘emotional’ and ‘proud’.

#### Experience of COVID-19

In the earlier posts, before testing commenced, many felt confident that they had COVID-19 stating that they ‘had textbook symptoms’ or ‘had seen enough cases to know’. Some had confirmation later in the year via an antibody test. Many reported receiving their COVID-19 vaccination.

#### Risk

Some GPs resigned themselves to ‘inevitably’ catching COVID-19. GPs commented that they were more vulnerable than other professions such as shop workers and those in secondary care, stating that 90% of patient contact is with GPs and contact is at closer proximity. Concerns around risk to BAME GPs centred around the disproportionately higher death rate in BAME GPs and calls for ‘appropriate measures’ to be put in place. These views were more commonplace among BAME GPs.

Once the vaccine was available, some GPs were frustrated by delays to their own vaccination.

In terms of concern for family members, some GPs talked about ‘living in fear of unknowingly passing it on to my family and loved ones’, particularly more vulnerable family members.

#### Communication

There was praise for primary care teams ‘pulling together’ and a clear sense of ‘solidarity’, alongside comments about how well community teams and volunteer/good neighbour schemes worked with GPs. While closer working relationships between primary and secondary care were referenced in some posts, there was a realisation that this was much needed. A ‘Berlin Wall’ and a ‘them and us mentality’ was described between primary and secondary care. Criticisms of hospital communications included hospitals ‘bouncing back’ GP referrals, and delays in patient test results. There were calls for better IT systems and for secondary care staff to spend time in primary care. Some praised technology and ‘online channels’ that enabled improved communication and made the situation more ‘bearable’.

Pleas were also made for better communication between the NHS and government, particularly as GPs had no warning of policy announcements such as shielding changes, and flu and COVID-19 vaccine roll-outs.

#### Self-care

GPs were very aware of the potential impact of the pandemic on their mental health; some reported looking after themselves, mostly through exercise and eating well, as well as some ‘self-care’ activities. The importance of taking annual leave and having days off ‘even in the middle of a pandemic’ was also emphasized.

Others, though, disliked the ‘self-care mantra’ and felt resilience planning was insufficient to ‘reverse the unprecedented levels of stress faced by primary care doctors today’.

## Discussion

The engagement of UK GPs’ with Twitter made it possible to conduct a mixed-methods social media analysis to explore large volumes of tweets relating to their perspectives and wellbeing during the pandemic and to compare this with pre-pandemic. The analysis reveals trends in the social commentaries made by GPs during the pandemic, including issues pertinent to GPs that may have affected their wellbeing. In our quantitative analysis, amongst a number of interesting patterns observed over this period, those of particular note relate to the strength of feeling around protection, risk and testing during the first wave of the pandemic; communication issues with and lack of appreciation by government, secondary care and the public, and peaks in commentary during the initial COVID-19 vaccine rollout. Similarly, our qualitative thematic analysis revealed key issues around perceived lack of resources and support, which had implications for GPs’ safety, workload and wellbeing. Perceived lack of support from government, media and the public affected their morale.

Our analysis identified comments about wellbeing which in 2019 were more predominantly related to patients, whereas 2020 saw more focus on GP wellbeing. Posts related to coping commonly reflected work pressures and fluctuated throughout the time period studied, suggesting that such views were prevalent well before the pandemic. Our qualitative findings highlight the perceived sources of increased workload and stress during the pandemic, including rapid moves to remote working (with remote consultations described as taking longer), GP self-isolation or shielding increasing pressures on colleagues, poor or confusing dissemination of policy guidance, increased patients with mental health problems, time taken cleaning and donning/doffing PPE. Meanwhile, frustrations were raised following media and even government references to GP practices being ‘closed’.

One other study has explored health care professionals’ wellbeing using social media, finding issues of lack of PPE and testing and changes in practice due to telemedicine predominate amongst US doctors [16]. Top phrases by physicians were ‘help us’ and ‘need PPE’. This concern was also voiced by UK GPs in our study. The US study also found discourse regarding unemployment (including furlough and pay cuts) was high among US physicians [16], which we did not identify. This may reflect differences between US and UK primary care.

It is already known that health professionals use social media to create virtual communities [15]. This is also evident from our study by how many of the GPs in our sample follow and send messages (often of support) to other GPs, including to others in our sample, demonstrating the degree of connectivity and support between GPs on Twitter.

## Strengths and limitations

The use of social media to explore GP wellbeing is novel and this paper is the first focusing on UK GPs. Our analysis ended in February 2021 but debate has continued, for example tensions arose in May 2021 due to a further call from NHS England and Improvement for GP practices to be ‘open’. GP workload resulting from public misconceptions about lateral flow testing for symptomatic patients was just beginning to be discussed in our data.

As with all social media research, we are limited by its content and by the sample. Online personas may be different to offline personas and GPs may be strategic in how and what they post. GPs have in the past been sued for discussing patients in such forums. Carville (2020) suggests they may also be cautious in discussing workplace issues for fear of disciplinary action [37]. GPs in our sample appeared to share their general views and opinions openly, but there was a tendency for them to refer to experiences and concerns around the wellbeing of colleagues, public access to GP services or public mental health rather than discussing their own personal experiences.

Twitter may not fully represent the demographics of the GP population. In general, social media users tend to be younger [38] and have a higher level of education [38-40]. In other respects, such as gender, race and ethnicity they tend to reflect the population [39, 40]. Although we identified GPs from different ethnic groups and regions, our sample was somewhat over-representative of GPs who are white, male, and living in London.

## Conclusions

Our analysis of the Twitter timelines of UK GPs indicates clear trends in the social commentary of GPs during the COVID-19 pandemic, some of which have implications for GP wellbeing. Discussion of perceived workload pressures, unsafe working practices, lack of support and abuse reflect wider media commentaries during this time. We have also demonstrated the value of collecting real world online data through informal social media posts, which is uniquely representative of lived experience across large samples.

Insights demonstrate the impact of the pandemic on existing pressures that may provide some potential areas for improvement in GP wellbeing: communication with secondary care, investment in staff and other resources, media relations and a need to re-establish teams and wider networks that were disrupted during the pandemic.

## Data Availability

Data are publicly available on Twitter. Due to ethical constraints and the terms and conditions of Twitter we are unable to provide direct access to the data.

## Ethical Approval

This study was approved by the Health Sciences Research Governance Committee, University of York in December 2020. No HRA approval was required for this study.

## Acknowledgements

We would like to thank Professor Mike Thelwall, creator of Mozdeh at the University of Warwick for providing the Twitter dataset used in this analysis, and to the members of our Project Steering Committee meeting for their contributions throughout the design and conduct of this study: Prof Dame Clare Gerada, Prof Michael West, Prof Michael Holmes, Prof Tim Doran, and our Patient and Public Representatives for their contributions; Patricia Thornton, Stephen Rogers and Emma Williams.

## Data Availability

Under the Terms and Conditions of Twitter and in the interests of anonymity of the GPs in our sample. We are unable to provide access to our data. However, the posts were all publicly available on Twitter.

