## Supplementary Material for "General Practitioner perspectives and wellbeing during the COVID-19 Pandemic: a mixed method social media analysis"

Supplementary Table 1: Top ten hashtags in 2019, 2020 and 2021

| Top hashtags according to the number of mentions among the 185 GPs in 2019, 2020, and 2021 | | | | | |
|---|---|---|---|---|---|
| Top 2019 Hashtags | No. of mentions 2019 | Top 2020 Hashtags | No. of mentions 2020 | Top 2021 Hashtags (Jan to 12th Feb only) | No.of mentions 2021 |
| #nhs | 470 | #covid19 | 1014 | #covid19 | 122 |
| #rcgpac | 201 | #nhs | 717 | #nhs | 76 |
| #teamgp | 183 | #coronavirus | 296 | #covidvaccine | 74 |
| #babylosshour | 165 | #teamgp | 192 | #teamgp | 59 |
| #gp | 144 | #covid_19 | 170 | #geribookclub | 41 |
| #brexit | 129 | #covid | 146 | #covidvaccination | 34 |
| #menshealth | 126 | #primarycare | 125 | #covid | 30 |
| #primarycarenetworks | 112 | #covid19uk | 124 | #oneteam | 24 |
| #sepsis | 89 | #gp | 122 | #medTwitter | 18 |
| #primarycare | 88 | #generalpractice | 98 | #primarycare | 16 |
| Top hashtags according to the number of GPs using them in 2019, 2020, and 2021 | | | | | |
| Hashtags | GPs mentioning hashtag in 2019 (n=185) | Hashtags | GPs mentioning hashtag in 2020 (n=185) | Hashtags | GPs mentioning hashtag in 2020 (n=185) |
| #nhs | 68 (37%) | #covid19 | 103 (56%) | #covid19 | 33 (18%) |
| #brexit | 32 (17%) | #nhs | 73 (39%) | #nhs | 29 (16%) |
| #teamgp | 31 (17%) | #covid_19 | 45 (24%) | #covidvaccine | 26 (14%) |
| #gp | 29 (16%) | #coronavirus | 41 (22%) | #teamgp | 19 (10%) |
| #rcgpac | 23 (12%) | #covid | 38 (21%) | #covidvaccination | 17 (9%) |
| #primarycare | 23 (12%) | #covid19uk | 37 (20%) | #covid | 12 (6%) |
| #generalpractice | 22 (12%) | #teamgp | 34 (18%) | #primarycare | 12 (6%) |
| #mentalhealth | 21 (11%) | #gp | 33 (18%) | #vaccine | 10 (5%) |
| #socialprescribing | 19 (10%) | #nhsheroes | 28 (15%) | #lockdown | 8 (4%) |
| #health | 18 (10%) | #socialdistancing | 27 (15%) | #covid19uk | 8 (4%) |

*Key*

Red =current affairs, Blue – health or health service, Orange = COVID-19, Green =primary care, Grey =miscellaneous

### Supplementary Table 2: Top ten handles in 2019, 2020 and 2021

| Top handles according to the number of mentions among the 185 GPs in 2019, 2020, and 2021 | | | | | |
|---|---|---|---|---|---|
| Handle | Times used 2019 | Handle | Times used 2020 | Handle | Times used 2021 |
| @rcgp | 1042 | @rcgp | 778 | @nikkikf | 160 |
| @lowcarbgp | 514 | @nikkikf | 634 | @nhsengland | 78 |
| @nikkikf | 492 | @drsdeg | 578 | @rcgp | 57 |
| @nhsengland | 423 | @nhsengland | 535 | @parthaskar | 53 |
| @helenrcgp | 419 | @matthancock | 407 | @drsimonhodes | 49 |
| @drsdeg | 406 | @fhussain73 | 390 | @trisha_the_doc | 49 |
| @matthancock | 385 | @thebma | 350 | @yvettedoc50 | 49 |
| @sonalikinra | 339 | @trishgreenhalgh | 345 | @rbkingston | 43 |
| @thebma | 290 | @lowcarbgp | 326 | @drjamesgill | 40 |
| @fhussain73 | 284 | @movemoresheff | 311 | @nhskingston_ | 39 |
| Top hashtags according to the number of GPs using them in 2019, 2020, and 2021 | | | | | |
| Handle | No of GPs 2019 (n=185) | Handles | No of GPs 2020 (n=185) | Handle | No of GPs 2021 (n=185) |
| @rcgp | 100 (54%) | @rcgp | 83 (45%) | @nikkikf | 42 (23%) |
| @matthancock | 73 (39%) | @nhsengland | 76 (41%) | @nhsengland | 33 (18%) |
| @nhsengland | 68 (37%) | @matthancock | 76 (41%) | @rcgp | 30 (16%) |
| @helenrcgp | 58 (31%) | @nikkikf | 75 (41%) | @drsdeg | 19 (10%) |
| @nikkikf | 50 (27%) | @drsdeg | 64 (35%) | @drmarkporter | 18 (10%) |
| @thebma | 49 (26%) | @trishgreenhalgh | 64 (64%) | @primarycarenhs | 17 (9%) |
| @drsdeg | 46 (25%) | @borisjohnson | 55 (30%) | @matthancock | 17 (9%) |
| @bmj_latest | 37 (20%) | @thebma | 53 (29%) | @fhussain73 | 16 (9%) |
| @borisjohnson | 36 (19%) | @phe_uk | 45 (24%) | @helenrsalisbury | 16 (9%) |
| @drgandalf52 | 35 (19%) | @martinrcgp | 43 (23%) | @thebma | 16 (9%) |

*Key*

Red =politicians, Blue – organisation, Green = GPs or heads of organisations, Grey =miscellaneous

### Supplementary Table 3: Top ten words used in 2019, 2020 and 2021 (excluding stop words)

| Top words according to the number of mentions among the 185 GPs in 2019, 2020, and 2021 | | | | | |
|---|---|---|---|---|---|
| Top words used in 2019 (n=185) | Times used | Top words used in 2020 (n=185) | Times used | Top words used in 2021 (n=185) | Times used |
| Thank | 2739 | Thank | 3585 | Thank | 507 |
| Great | 2385 | Time | 2405 | Vaccine | 366 |
| GP | 1998 | Great | 2213 | patient | 332 |
| Patient | 1935 | Patient | 2191 | Time | 293 |
| Time | 1924 | People | 2190 | People | 290 |
| Good | 1747 | Good | 2128 | Great | 258 |
| Work | 1735 | Work | 2107 | Covid | 252 |
| People | 1460 | GP | 2013 | GP | 249 |
| care | 1368 | Care | 1791 | Good | 248 |
| Health | 1226 | Health | 1394 | Work | 247 |

| Top words according to the number of GPs using them in 2019, 2020, and 2021 | | | | | |
|---|---|---|---|---|---|
| Top words used in 2019 (n=185) | No of GPs 2019 (n=185) | Top words used in 2020 (n=185) | No of GPs 2020 (n=185) | Top words used in 2021 (n=185) | No of GPs 2021 (n=185) |
| Thank | 153 | Thank | 154 | Thank | 154 |
| Great | 152 | Time | 147 | Time | 147 |
| Work | 146 | Great | 146 | Great | 146 |
| Time | 142 | Work | 146 | Work | 146 |
| Good | 141 | People | 145 | People | 145 |
| Health | 140 | Good | 141 | Good | 141 |
| Patient | 136 | Patient | 139 | Patient | 139 |
| People | 135 | Please | 137 | Please | 137 |
| Care | 131 | GP | 137 | GP | 137 |
| GP | 131 | Health | 135 | Health | 135 |

*Key*

Blue – health or health service, Orange = COVID-19, Green =primary care, Grey =miscellaneous

### Supplementary Table 4: Trends in terms used for selected topics 2019 to February 2021

| Topic | Terms Searched | Highest Percentage of Daily Tweets | Notes |
|---|---|---|---|
| **Vaccination** | Vaccination OR vaccinations OR vaccine OR vaccines OR vaccinated OR vaccinating OR #covidvaccine OR #vaccines OR #covidvaccination OR#vaccineswork OR #covid-19 OR#covid2019 OR #vaccine OR #vaccination OR #covidvaccines OR #covid19vaccine OR #vaccinessavelives OR #pfizervaccine OR #pfizer OR #astrazeneca OR #covidvacccine #vaccinate OR #getvaccinated | 23.4% | Prevalent from the 9th November 2020 onwards, with high volume from beginning of December. 9th November 2020 Pfizer announced results of their phase 3 clinical trial efficacy test results. Vaccination roll out began on 8th December 2020. |
| COVID-19 | corona OR coronavirus OR covid OR covid19 OR pandemic OR virus#covid19 OR #coronavirus OR #covid2019 OR #covid_19 OR #covid OR #covid_19 OR #covid19uk OR #coronavirusuk OR #covid-19 OR #fightcovid19 OR#pandemic OR #covid__19 OR #corona OR #covid_19uk OR #coronavirusoutbreak OR #coronavirusupdate OR#covid2019uk OR #covid19pandemic OR #longcovid OR #covidassessmentcentre OR #coronacrisis OR #coronaviruspandemic OR #coviduk OR #coronacrisisuk OR #zerocovid OR #coronauk OR #coronavirusupdates OR#coronaoutbreak OR #coronaupdate OR #coronvirusuk OR #covidiots | 18.1% | Prevalent from Feb and March 2020 onwards. COVID became notifiable disease in UK on 5th March 2020. |
| Interventions to reduce COVID-19 transmission | distancing OR isolate OR isolated OR isolating OR isolation OR lockdown OR mask OR socially OR transmission OR transmitted OR transmit OR vulnerable OR wave OR shielding OR #uklockdown OR #stayathomeandstaysafe OR #stayhomesavelives OR #lockdown OR #lockdown2 OR #lockdownuk OR #lockdownnow OR #socialdistancing OR #socialdistancinguk OR #stayhome OR #stayathome OR #staysafe OR #wearamask OR #flattenthecurve OR #stayinworkout OR #covidiots OR #washyourhands OR #wearamask OR #physicaldistancing OR #savelives OR #stayathomesavelives OR #stayhomestaysafe OR #mask OR #facemask OR #shielding OR #mymaskprotectsyou OR #staysafestayhome OR #selfisolation OR #flattenthecuve OR #stayathomeprotectthenhssavelives OR #protectthenhs OR #facemasks OR #saveournhs OR #coronaviruslockdownuk OR #selfisolating OR #socialdistancinguk OR #covidiot OR #mask OR #selfisolate OR #trackandtrace OR #coronaviruslockdown OR #socialdistanacing OR #shielding OR #coronalockdown OR #coronaviruslockdown OR #coronalockdown OR | 21.8% | Prevalent from March 2020 onwards.<br>Peaks on topics of lockdowns, Dominic Cummings, and mask campaigning<br>23 March 2020, the UK went into lockdown.<br>22 May News breaks of Mr Cummings trip to Durham, 24 July Compulsory face coverings in indoor public spaces, 31 October 2020 announcement of second lockdown |

| | | | |
|---|---|---|---|
| | #social_distancing OR #socialdistance OR #stayalert OR #keepyourdistance OR #isolation | | |
| **Remote Working** | Telephone OR phone OR video OR OR virtual OR remote OR remotely OR teleconsultation OR accurx OR footfall OR econsult OR skype OR zoom OR #remoteworking OR #accurx OR #remoteconsultations OR #doctorlink OR #onlineconsultations #econsultations OR #teleconsultations OR #virtualgroupconsultations OR #newmodelsofconsultation OR #newconsultationmodels OR #videoconsultation OR #virtualconsulting OR #virtualmedicalmeetings OR #virtualmeetings OR #zoom OR #saturdayzooming OR #zoomlife OR #workingfromhome | 14.2% | Prevalent throughout 2019 to 2021. More consistently discussed from February 2020 onwards. Slight upward peak in March 2020 and larger peak 30 July 2020.<br>NHS Long Term Plan committed practices to offer e-consultations from April 2020.<br>30 July 2020 Matt Hancock state GPs 'should do all consultations remotely going forward'. |
| **Testing** | test OR tested OR testing OR #covidtesting OR #testhealthcareworkersnow OR #testnhsstaff OR #testtesttest OR #testingforcovid19 OR #testthenhs OR #coronavirustesting OR #testing | 12.9% | Prevalent from March 2020 onwards<br>Peak in first wave and September 2020<br>Testing was not widely available during the first wave even for HCPs.<br>Shortages in testing was reported in September with people unable to get a test or travelling for miles to get one. |
| Safety | at-risk OR danger OR dangerous OR dangerously OR safe OR safely OR safer OR safety OR scary OR scare OR scaremongering OR risk OR risking OR risky OR unsafe | 11.1% | Prevalent from March 2020 onwards<br>Peak in March –April 2020<br>First COVID death initially reported on 5 March 2020 (later earlier death on 30 January 2020 identified as first death).<br>25 March 2020 - first GP death. PPE shortages were widely reported during the first wave. |
| **Coping** | burnout OR cope OR coping OR difficult OR distress OR distressed OR distressing OR resilience OR stress OR stressed OR stressful OR struggle OR struggled OR struggling OR tough OR #burnout OR #resilience OR #physicianburnout OR #tackleburnout OR #coping OR #nhsresilience OR #betterworkloadnotmoreresilience OR #lessworknotmoreresilience OR #stress OR #workstress OR #doctorsindistress OR #stressed | 9.6% | Fluctuates throughout 2019 to 2021. Peaks reflect increase in posts on work related stress. |
| PPE | ppe OR protect OR protected OR protecting OR protective OR apron OR aprons OR glove OR gloves OR gown OR gowns OR doff OR doffing OR don OR visor OR visors OR goggles OR n95 OR ffp2 OR ffp3 OR #PPE OR #ppefornhs OR #ppeshortage OR #ppenow OR #properppe OR #weneedppe OR #ffp3 | 9.6% | Prevalent from March 2020 onwards. Peak on the 22nd April 2019.<br>PPE shortages were widely reported during the first wave. |
| **Staff** | frontline OR staff OR frontliner OR #nhsheroes OR #frontlineheroes OR #frontliners OR #nhscovidheroes | 9.1% | Peaked in March to April 2020 with discussion of PPE and testing of staff and December 2020 to January 2021 with vaccinating staff. |
| **Mortality** | death OR die OR died OR mortality OR #death OR #mortality | 8.0% | Peak in first wave |

| | | | |
|---|---|---|---|
| | | | First COVID death initially reported on 5 March 2020 (later earlier death on 30 January 2020 identified as first death).<br>25 March 2020 - first GP death. |
| **Sadness** | sad OR sadly OR sadness OR #sadly | 6.3% | Fluctuates throughout 2019 to 2021 |
| **Enjoyment** | enjoy OR enjoyable OR enjoyed OR enjoying OR enjoyment OR joy OR #joy | 6.3% | Dipped after the end of January 2020. Often referred to enjoying work, a course or learning. Before start of the pandemic more about enjoying an event such as cinema, xmas and after pandemic began walks, runs, cycling or countryside |
| **Appreciation** | appreciate OR appreciated OR grateful OR #clapforourcarers OR #gratitude OR #clapforcarers OR #clapforthenhs OR #grateful | 6.3% | Fluctuates throughout 2019 to 2021 |
| **Wellbeing** | wellbeing OR well-being OR #wellbeing OR #mindfulness OR #wellness OR #nhswellbeing OR #wellbeingatwork OR #healthandwellbeing OR #worklifebalance OR #mindfullness OR #wellbeingmatters OR #worklife OR #healthandwellness OR #doctorwellbeing OR #nhsmentalwellbeing OR #waystowellbeing OR #bmawellbeing OR #staffwellbeing OR #juniordoctorwellbeing OR #workplacewellbeing OR #clinicianwellbeing OR #worklifebalance | 5.6% | More peaks before COVID-19 in March 2020. Often refers to support available, particularly for GPS after start of pandemic and for patients and staff before. |
| **Party** | Party | 4.6% | More prevalent more throughout 2019 than 2021. Mostly political parties mentioned.<br>UK general election 12 December 2019. |
| **Overworked** | overstretched OR over-stretched OR overwhelmed OR overwhelming OR overworked OR relentless OR relentlessly OR stretched OR unrelenting OR work-life OR workload OR work-load OR #gpworkload OR #workload OR #workloadpressures | 4.6% | Peaks on 23 January 2019 discussion on GP pharmacist role in reducing GP workload.<br>5th September 2020 discussion on workload and F2F.<br>18th October 2020 discussion on abuse of primary care<br>7th January 2021 talk of workload prioritisation and vaccines 18th |
| **Closed** | closing OR closed OR close OR #gpsareopen OR #gpisopen #weareopen OR #generalpracticeisopen | 4.3% | More frequently used in September 2020.<br>Early September 2020 NHS England wrote to all GP practices to ‘reopen’ |
| **Flu** | Flu OR #flu OR #flujab OR #fluvaccine OR #influenza OR #fluclinic OR #flujabs OR #flu2020 OR #getyourflujab OR #flu2019 | 4.2% | More prevalent from September to November 2020 and in January 2019.<br>1 September 2020 start of largest UK flu vaccination programme |
| **F2F** | F2F | 3.2% | More frequently used after March 2020. |

| | | | |
|---|---|---|---|
| | | | NHS Long Term Plan committed practices to offer e-consultations from April 2020 |
| **Morale** | demoralised OR demoralising OR demoralise OR morale OR #morale | 2.5% | More frequently used in September 2020.<br>September 2020: Media coverage suggesting GP surgeries have been closed |
| **Incompetence** | incompetence OR incompetent OR #incompetent | 2.2% | Prevalent more from April 2020 onwards. Discussions around government handling of pandemic<br>Media reports suggested government 'incompetence' increased threat of COVID-19 |
| **Bashing** | bashing OR bashed OR bash OR abuse OR abused | 1.3% | More frequently used in September 2020. GP abuse or bashing common thread in September 2020.<br>September 2020: Media coverage suggesting GP surgeries have been closed |

### Supplementary Table 5: Main Category, explanation and examples

| **Category** | **Explanation** | **Paraphrased examples from multiple GPs*** |
|---|---|---|
| Changes to GP practice | Includes changes in practice due to COVID such as with video and phone consultations (poor internet, poor reception, communication difficulties), working from home, and adaptions with COVID related work.<br>Does not include increased workload. | ***Challenges to practice (negative)***<br>*55% of communication is non-verbal (body language). So just think what GPs might be missing with the transition to using online consultations and texts to communicate with patients?*<br>*Not everyone wants a remote consultation. It's limited by Internet connections and exposes the digital divide and people do not seem to be ready to pick the phone up at 8:30am!*<br>*From personal experience, remote consultations are more exhausting than F2F ones. Putting us at a higher risk of burnout.*<br>*Never more challenging time to help patients in these times of distancing. Really missing face to face interactions and human connections that were integral before all this.*<br>*Good Morning? My GP clinic should have started 30 minutes ago but I'm still rebooting, because nothing will load*<br>*Clinics can run late for a variety of reasons. Some patients need the extra time to be helped. Emergencies come in. But running late because of IT is not good!*<br>*Given a decade of cuts and still we deliver. Serco get 12 billion and we get £12.58 per vaccine. Where will we put patients to wait 15 mins after their jab, adequately socially distanced? How will we also do our day jobs?*<br><br>***Changes to practice (positive or neutral)*** |

| | | |
|---|---|---|
| | | *I was worried about how I'd navigate the changes in GPland during COVID however I am pleasantly surprised at how much better the systems are now! All patients seen today through ways that worked for them - face to face, telephone, video consultations! Happy patients = happy GP*<br>*Amazing achievement on how quickly and how brilliantly primary care adapted to remote triage and working. Things we'd been trying to achieve forever happened almost overnight.*<br>*I've found remote consults really helpful and often more efficient. Hope we can take the best of these changes into the future!* |
| NHS resources | Refers to funding of the NHS and general practice in terms of resources (such as funding, staff, PPE, and C19 testing).<br>Does not to include resources unrelated to GPs such as ventilators (unless has an impact on GPs or GP work). | ***Lack of PPE***<br>*If we are going to get through #covid19 we need the appropriate PPE and testing. Urgent action required now!*<br>*Still no proper PPE in general practice. I'm so livid. I'm putting myself and family at risk because of this shambles government. Sort it out and sort it out fast.*<br>*I understand why hospital staff are getting PPE but why not GPs who are in the frontline dealing with many more people.*<br>***Lack of COVID-19 Testing***<br>*We need testing urgently. Almost half of our GPs are now off in our practice because of household isolation. If primary healthcare collapses there will be even more pressure on hospitals.*<br>*Test GPs for #COVID19 so that we can get on with our jobs without putting the public at risk*<br>*Prince Charles, politicians, premiership footballers and the super-rich are all getting tested for coronavirus yet GPs like me who are on the front line are being told no.*<br>*Don't forget GPs! As a GP I'm not yet categorised as a 'priority' for getting testing, despite the fact GPs provide patient-facing out of hours and urgent care throughout pandemic. So with no test I'm at home, my shifts cancelled and patients suffering. This is not right*<br>*It took me 10 days (supposed to be 2!) to get my results - in the meantime I can't see my patients face to face. This is not by choice, but compulsion owing to a failing system.*<br>***Lack of Staff***<br>*All this with a 1000 fewer GPs and 400,000 more patients than last year! More work, with fewer GPs, under the most challenging circumstances ever faced by the #NHS - that sums up #GeneralPractice right now.*<br>*We are losing GPs at an ever alarming rate. Our workforce is overstretched and underfunded with extremely low morale and facing constant attacks from the media and public.*<br>*Seeing the 'save the NHS' narrative replaced with the customary GP bashing at a time when we have fewer GPs than in 2005 is only going to push more of us away.*<br>***Lack of Funding or Pay***<br>*Government was right to give support to businesses for staying closed, but why is that not being as readily given to GP practices for staying open? We spent ££££s of our OWN money on PPE, and on locum GPs so we could provid a full service throughout the pandemic.* |

| | | |
|---|---|---|
| | | *The totally shocking thing is that the failed #SercoTestAndTrace system costs MORE than ALL the GPs in the whole country for a whole year. Just think about what primary care could do with that money.*<br>*Some doctors who die during this pandemic will not receive full NHS death benefits. At a time when doctors are putting themselves in harms way the govt must act.*<br>*I hope that I'm wrong but it seems that nurses, healthcare assistants and foundation doctors will not benefit from a pay rise... Why ever not?!*<br>***Lack of resources (general)***<br>*We're working hard to do our best for patients with very limited resources. I don't know any doctors who voted for this threadbare and starved NHS.*<br>*GPs need support to do their job and for patients. Otherwise hospitals will be overwhelmed and prioritising sick patients will be become even harder.*<br>*I do try not to let myself get angry as I know it won't help me. But I am so so angry about the decade of cuts and how GPs will be left to make do and mend.*<br>***Adequate Resources***<br>*Good news! GP surgeries will be provided with PPE from this week free of charge.*<br>*Whilst it feels odd as a GP dressed in full PPE I am feeling safer and better protected.*<br>*So lucky to have received donations of masks, visors and gloves. Lately, PPE supplies from the CCG have also been adequate.* |
| Direction/ Management/ Leadership from UK government or leading organizations such as RCGPs and BMA | This could be positive or negative opinion regarding the management of the NHS and/or management of the COVID crisis that impacts on GPs directly or indirectly.<br>This excluded any tweets about general annoyance with government decisions, that could be shared by general public, and concentrated on comments relating specifically to an impact on GP working lives and wellbeing. | ***Direction, management (negative)***<br>*One huge problem is that when politicians talk about 'the NHS' they actually mean 'hospitals' and almost totally ignore the importance of primary care and public health.*<br>*I reiterated the "stick to the rules" mantra. @MattHancock - your teammates have made a farce of this while my teammates continue trying to save the lives you risk! #teamGP*<br>*Yet again @MattHancock prioritising headlines. Telling those aged 50-65 to get a flu jab with their GP without telling us first. Where are the vaccination fairies? Tomorrow is going to be chaos at our practice.*<br>*It is great the UK are going to start COVID vaccinations. Only problem, the GPs who are supposed to be organising this haven't been told anything about it!*<br>*Patients have been able to see their GPs face to face if needed throughout the pandemic. Stop blaming GPs and start addressing the failing test and trace system.*<br>*"Digital exclusion!!!!!!"... @MattHancock shows he has very little understanding about the vital role GPs play in the lives (+deaths) of so many. Remote consultations are NOT best for everyone and I will not accept his "instruction" for this to become the default option.*<br>*GPs have the infrastructure, knowledge and above all trust to run the test and trace system. We are already the gatekeepers and could provide a highly localised and nuanced system at a fraction of the cost of SERCO.* |

| | | |
|---|---|---|
| | | *Today tens of thousands are getting their flu jabs from GP practices. It will be safe and expertly well organised and without the need to travel far. Just think how different test and trace could have been if this government invested in primary care not a private company.*<br>*Call me cynical but the delivery of the first covid vaccinations in hospitals is not about logistics but a carefully staged photo opportunity and an insult to us in primary care, given GPs and practice nurses will be deliver the majority of the vaccines.*<br>***Direction, management (positive)***<br>*I'm so pleased the government is rolling out more testing. I want to see them ensuring that all healthcare professionals have access to testing.*<br>*It is a challenging time for everyone and cool heads are required. Thank you to all at the BMA for all the work you continue to do.*<br>*I don't know how @NikkiKF does it. So much respect for her doing an awfully hard job in difficult times.* |
| Information | This is misinformation, information use and sharing and information to support GP wellbeing.<br>Misinformation could be about GPs or GP surgeries (such as GPs closed or not seeing patients) or misinformation that impacts on GPs or GP work (such as mistakes in the patient shielding list or misinformation on when the public should contact a GP). Misinformation also included lack of information and information seeking (such as asking questions).<br>Misinformation is related to GPs or GP work not general misinformation to public. | ***Misinformation (about or received by GPs)***<br>*GP surgeries are not closed. GPs have continued to prioritise care in extremely difficult circumstances to make sure that people receive the care they need – frightening the public, insulting general practice and perpetuating false rumours is dangerous. Please stop this now!*<br>*Why are we working with version 9?? Who's has the energy to keep cleaning shielding lists? Why are patients told one moment they on and the next moment they're off the list? I'm am so livid with the whole process.*<br>*Who is writing prescriptions? I know of no GP surgeries that have close; except the ones that had to close well before the Covid due to lack of funding. I'm doing over 50hrs a week as are my colleagues. We are working remotely and getting patients in for face to face consults when deemed necessary. This is for everyone's safety.*<br>*So often in this pandemic my patients apologise for wasting my time. You are not wasting my time! We are here to care for you when you're ill and to assess and reassure if you are not - COVID or no COVID! We care and we are open!*<br>***Information use and sharing (among GPs)***<br>*Watch this walkthrough guide on how @accuRx can help your practice, improve patient safety, reduce workload and save you money*<br>*THIS is the best practical guide I have read to date regarding C19 in GPland. Coronavirus disease 2019 (covid-19): a guide for UK GPs*<br>*GPs and primary care sharing top tips and helping colleagues to cope with the challenge of rapid change due to COVID. Use #UKGPC19 for sharing ideas and good practice*<br>*Any GPs found useful resources to share with patients to teach them how to take their own pulse / respiratory rate?*<br>*I'm receiving lots of requests for sick notes for vulnerable/high risk categories patients whose employers are wanting them to continue working. Is there official guidance on this?*<br>***Information to support GP wellbeing***<br>*Many NHS staff are tired and covid weary. Join us to hear how to maximise your breaks, long or short, re-energise and reboot to help cope with what is to come. For ALL NHS staff* |

| | | |
|---|---|---|
| | | *Today I am really pleased to welcome and open @theBMA wellbeing support group, bringing together many organisations and people with the collective aim to support the wellbeing of doctors at this very trying time.* |
| Appreciation | Some form of recognition or lack of recognition for GPs work and sacrifices. Also GPs being listened to and opinions valued or being undervalued and the target of mistreatment such as abuse or theft. Appreciation could be from Government, general public, colleagues, family, friends, neighbours, businesses. GP appreciation of others is also included here. | ***Appreciation of GPs (negative)***<br>*We will continue to deliver the unprecedented flu vax programme to protect the most vulnerable during a very difficult winter. We will do this with the backdrop of continuing criticism from some, whilst demoralised and exhausted.*<br>*The unrelenting workload and constant criticism is pushing my colleagues away.*<br>*Seeing the 'supporting the NHS' narrative replaced with the customary GP bashing by some at a time when we have fewer GPs than we did in 2005 is only going to push more of my colleagues away.*<br>***Appreciation of GPs (positive)***<br>*Thanks for clapping! This gesture is massively appreciated by this GP! Now please put pressure on the government for all frontline staff to have access to adequate PPE and testing.*<br>*Clapping, praises and thanks are appreciated by GPs but what we really needed is responsible behaviour, and consideration from the public.*<br>***Appreciation of others***<br>*To my lovely neighbours who left me this, thank you! Whilst this is understandably an anxious time for everyone, look out for each other, be kind to others, especially to vulnerable people.*<br>*Thank you to the bike shop for fixing my broken bike as part of a free NHS staff bike service...safe to cycle to work again tomorrow!*<br>*I cannot justify in my head why I should get special priority shopping when I'm just doing my job (which I love and am doubly grateful to still have in these difficult times).*<br>*GPs throughout the country are working incredibly hard under very difficult circumstances and an acknowledgment of that by politicians on the BBC today is refreshing compared to the attacks we have received recently.* |
| NHS work colleagues | Refers to work of NHS work colleagues, either in primary care, community care, secondary care or management. | ***NHS work, colleagues (positive)***<br>*Really happy to invite the BBC into my GP practice when I was working today. My clinical and admin colleagues are amazing champions of the flu vaccination programme!*<br>*Not all heroes wear capes but recently it has become evident that many of them wear NHS badges*<br>***NHS work, colleagues (negative)***<br>*I have the surreal feeling that what goes on in the NHS management centre and what goes on on the front line are operating in two separate universes.*<br>*Reviewed 10 letters from hospital colleagues stating 'GP refused to see', yet these patients all had no contact with our practice at all. This needs to be improved.*<br>*In GP land we have electronic transmission of prescriptions to community pharmacies. This has been fantastic during Covid. Why can't this be extended to secondary care?*<br>*I am a GP and I disagree with GP bashing (obviously) but in primary care we do not always work welltogether either.* |
| Personal GP experiences | These posts provide personal information about the tweeter (i.e. the | ***Experience C19 positive test/self-isolation*** |

| | | |
|---|---|---|
| | GP) themselves. For example a declaration of a positive COVID test or having the COVID vaccine. In addition personal feelings related to burnout or stress (such as overwhelmed, feelings that they cannot cope or mental/emotional exhaustion.<br>Other feelings such as frustration, anger, worry, sadness, grateful, proud, tired, or other experience (e.g. physical exhaustion). | *As of Sunday morning I started with a continuous dry cough, so have begun self-isolating. I wish I could be tested so that if negative I could resume work and my whole household wouldn't have to stay indoors for 14 days.*<br>*Happy and proud to have got my first dose of the Covid Vaccine. I feel great. Excellent work by the vaccination team*<br>***Experience burnout/stress***<br>*Completely exhausted and nearly broken but proud that I am part of an amazing team providing a fast and great response in adversity.*<br>*Feeling teary today. Every shift is harder than the last one.*<br>*Heartbroken. Mentally drained. We are living in a bad dream.*<br>*I can feel myself heading towards burnout.*<br>***Experience other feelings***<br>*Knackered and scared, but I will do carrying on doing my best as our patients deserve it.*<br>*If you are a HCP and haven't shed a few tears this week you probably don't appreciate what lies ahead.*<br>*Politicians are not listening to GPs or any signals from general practice about what is happening right now. I am living through this. I am frustrated and scared for my patients.*<br>*I feel guilty, frustrated and terribly miss being able to clinically examine a patient but I am shielding.*<br>*Tomorrow, I will be helping deliver the biggest vaccination program ever I'm proud to be part of this on such a historic day.* |
| GP workload | This may be a general statement about increasing or decreasing workload. For example, increasing workload due to changing practices due to COVID (such as remote consultations and additional cleaning). | ***Workload increase***<br>*We are carrying out more consultations per day than before COVID despite the longer consultations times. But we cannot sustain this much longer. Tired now. Low morale. And press and public alleging we are closed sitting with our feet up!*<br>*It is not just about numbers - in most instances face to face consults are much quicker compared to video consults. The biggest challenge is the increasing workload, it just feels relentless*<br>*Measuring F2F appointments as a proxy for service delivered in general practice makes no sense at a time when we are trying to protect patients by remote consulting. It is an incredibly challenging and busy time.*<br>*I am worried about the increasing number of patients broken with mental health problems, stress and anxiety. They need longer than 10 minutes, and even longer still when you are trying to support them remotely.*<br>*Please DO NOT phone GP surgeries trying to get a Covid Vaccine We are inundated with calls and requests whilst trying to do our normal work.*<br>*It is not unusual to be booked until 2200 hrs with a list of econsults and tel triage that starts at 8.30 am at 10min intervals.*<br>*TEN MINUTES?? I can just about manage some of the time before covid. How can we work to 10mins face to face when you have all the extra faffing to do with donning/doffing/disinfecting chairs, couch and equipment in between patients?*<br>***Workload decrease***<br>*Normally this is an incredibly busy at this time. This is so strange.*<br>*Back in April there were 2 to 3 weeks that were very quiet very eerie.* |

| | | |
|---|---|---|
| Colleagues health or wellbeing | This could be a general statement about anxiety, depression or burnout or COVID illness or death among NHS colleagues or family working in general practice. | *Concern about colleague health, wellbeing (depression, burnout, etc.)*<br>*In primary care the mental and emotional health of our staff is being affected by increasing complaints and verbal abuse from a small but significant minority of patients.*<br>*Our workforce is exhausted, burnt out, isolated or sick.*<br>*Seen so many colleagues suffering during this pandemic with anxiety and stress. We need to pull together.* |
| Risks to GPS | This is in the form of the dangers of the occupation in terms of catching COVID and death. This can be to the GP themselves and their families to whom they may transmit it. | ***Risks to GPs themselves***<br>*GPs are highly vulnerable as we see many ill patients every single day. We could be super spreaders and should be given priority testing and adequate PPE*<br>*Whilst we in primary care work our absolute hardest, and risk our lives to help others against COVID it's so insulting to see people going on nights out, ignoring the seriousness of it all and carrying on as normal. If we can risk our lives and stay at work for you, you can get bored and stay home for us.*<br>*I risk my life every day and all I have to look forward to is tax increases, exhaustion, potential death and a clap every Thursday night.*<br>*Not sure the staff in supermarkets examine many earholes or anuses or throats or vaginas or skin rashes or scalps or testicles and therefore tend not get as close and personal with their customers. Maybe I'm missing something here. Bias or bitterness?*<br>*This is sadly yet another example of the perception that Covid is only a secondary care matter. We see more patients than our colleagues in secondary care, is it's not conceivable that our exposure is at least as high?*<br>***Risks to GPs families***<br>*Our 5 year old thinks I don't love him anymore because I daren't get too close. How do I explain social distancing to the children of the unprotected, untested NHS front liners!*<br>***Risks to BAME GPs***<br>*In the UK 12 GPs have died from COVID-19 and 11 were BAME*<br>*We need appropriate measures in place to protect BAME colleagues who are more at risk* |
| Communication | Communication, integration or collaboration between primary care and other NHS departments or sectors such as secondary care. | ***Communication (positive)***<br>*Brilliant to see the NHS PrimaryCare and SDEC working together to manage COVID19 in the community.*<br>*Really proud of the collaborative work being done to support the Covid vaccine delivery. Communication at its absolute best.*<br>***Communication (negative)***<br>*The pandemic has highlighted the long-overdue need to knock down the Berlin wall between primary and secondary care that works against patient care and HCP stress and job satisfaction.*<br>*I would never pass judgement on hospital staff workload especially on social media. Most hospital doctors have never done a GP rotation. So why do some think it's ok to pass judgement on GPs?* |

| | | |
|---|---|---|
| Self-care of GPs in reference to their wellbeing | This could be taking action to help with their wellbeing or work-life balance. For example, going on a course on wellbeing, or taking a walk to relieve the stress. This needs to be explicitly stated to be related to their wellbeing. Activities such as watching football or going for a walk with no mention of impact on wellbeing were excluded. | ***Self-care (positive)***<br>*We need to maximise all self-care and kindness just for us to survive this intact.*<br>*Don't forget to take care of yourself both mentally and physically. This is a difficult time for many. I have been taking my own advice and keeping a diary of my thoughts and feelings. It does helps..*<br>*Busy couple of days covering Annual leave. It is so important to take a break to protect wellbeing even in the middle of a pandemic.*<br>***Self-care (negative)***<br>*No amount of self-care or resilience training will reverse the unprecedented levels of stress faced by general practitioners in the NHS today.* |

*Quotations were paraphrased to anonymise individuals, while still providing examples of commentary by theme. No more than two quotes were selected from the same GP.
